# PAUL^®^ glaucoma implant – efficacy and safety: A systematic review and meta-analysis

**DOI:** 10.1101/2025.11.01.25339321

**Authors:** Chuen Yen Hong, Aaron BC Wong, Chuen Lin Hong

## Abstract

**Background:** This systematic review and meta-analysis aim to evaluate the efficacy and safety of the PAUL^®^ Glaucoma Implant in reducing intraocular pressure and number of anti-glaucoma medication use in both adult and paediatric populations.

**Methods:** A systematic review was conducted following the PRISMA guidelines. The databases PubMed, Ovid-Embase and Scopus were searched to include studies published between January 2017 and September 2025. Studies were stratified by age and risk of bias was assessed using the Newcastle-Ottawa Scale for observational studies and ROBUST-RCT for randomized trials. Primary outcomes were intraocular pressures and number of glaucoma medications at each follow-up visits. Random effects meta-analyses were performed. Secondary outcome was complications.

**Results:** Twenty-four studies (twenty adults, four paediatric) comprising 836 eyes were included in our review and meta-analysis. Both adult and paediatric patients showed significant IOP reduction post-surgery, with a mean difference of 17.48 mmHg (95% CI: 14.59, 20.37) and 20.31 mmHg (95% CI: 7.80, 32.82) at 1 week, respectively, and sustained reductions at 12 months. The reduction in glaucoma medications was 79.2% for adults and 71.4% for children at 12 months. Subgroup analyses demonstrated greater IOP reduction in studies conducted in the Middle East. The uses of Mitomycin C did not significantly affect outcomes.

**Conclusion:** The PAUL^®^ glaucoma implant showed significant and sustained IOP reductions with reduced need for glaucoma medications. The PAUL^®^ glaucoma implant is a promising surgical option for glaucoma management in both adults and children. Further long-term prospective comparative studies are needed to assess long-term efficacies and allow direct comparisons with other glaucoma drainage devices.

## Introduction

Glaucoma remains the major cause of blindness globally, accounting for 8.39% of all blindness.^1^ Intraocular pressure (IOP) is the only known modifiable risk factor, with the risk of primary open angle glaucoma (POAG) increasing by 1.13 times for every one mmHg increase in IOP.^2^ However, glaucoma occurs across various levels of IOP and diagnosis is not limited to those with elevated IOP. It is well established that lowering of IOP slows glaucoma damage regardless of whether baseline IOP is normal or elevated.^3–5^ Treatments either medical, laser, or surgical are all aimed at lowering IOP either by reducing the production of aqueous humour, increasing the outflow or both.

Glaucoma drainage devices (GDD) are widely used when patients have failed medical therapy, laser and previous trabeculectomy. Various devices have been developed since the introduction of the Molteno3 (Molteno Ophthalmic Limited, Dunedin, New Zealand) in 1966. In Australia and New Zealand, A 2020 survey of glaucoma surgeons reported that the most used GDDs was the Baerveldt glaucoma implant (BGI; Abbott Laboratories Inc., Abbott Park, IL).^6^ In 2017, clinical trials for the PAUL Glaucoma Implant (PGI; Advanced Ophthalmic Innovations, Singapore, Republic of Singapore) commenced. The PGI was a novel non-valved GDD designed to have a smaller internal (0.127mm) and overall (0.467mm) tube calibre compared to currently available devices with the aim to reduce risk of early post-operative hypotony.^7^

Recognising the increase in the use of PGI in clinical practice, we performed a systematic review and meta-analysis of the literature. Our aim was to summarise and assess the efficacy and safety of PGI in reducing IOP.

## Methods

The systematic review was conducted according to the Preferred Reporting Items for Systematic Reviews and Meta-Analyses (PRISMA) guidelines. The PRISMA checklist is shown in Supplementary 1.

### Search strategy

We performed a systematic review in 3 databases: PubMed, Ovid, and Scopus, from January 2017 till September 9, 2025. The search was limited to studies after 2017 as clinical trials for PGI only commenced in 2017. We also searched the reference lists of any article included after full-text review. There was no restriction on language. The detailed search strategies for each database are provided in Supplementary 2.

### Study selection

All articles retrieved was collated into a Microsoft Excel (version 2501; Microsoft, Redmond, Washington) sheet and duplicates were removed. The resulting list of titles and abstracts were independently screened by two authors (C.Y.H and C.L.H). Any article selected by at least one reviewer were reviewed in full for inclusion, with any discrepancies between the two reviewers resolved by discussion.

Studies were selected based on the following inclusion criteria: (a) original study of any design, (b) the study investigated PGI implantation, and (c) outcomes of interest were reported, including IOP reduction, IOP-lowering medications reduction and success rates. There were no restrictions on language, age or country.

We excluded review articles, letters, commentaries or editorials. Studies with incomplete or duplicated data were also excluded.

### Data abstraction and analysis

Study characteristics extracted included authors, publication year, study design, country of study, sample size, aetiologies included in the study, success criteria and follow-up duration. Data for patient demographics (and their baseline characteristics), IOP measurements, number of IOP-lowering medications, success rates, complications and adverse events were abstracted in Microsoft Excel (version 2501; Microsoft, Redmond, Washington). Studies were stratified into adult and paediatric populations.

### Quality assessment

The risk of bias of the included observational studies was assessed using the Newcastle-Ottawa Scale^8^ and ROBUST-RCT^9^ for randomised controlled trials, rated by two independent reviewers (C.Y.H and C.L.H) with disputes resolved by discussion. Details are in Supplementary 3.

### Statistical analysis

The primary outcome of interest was mean IOP and glaucoma medication reduction. Secondary outcome was the number (proportions) of eyes with early (< 3 months post- operative) and late complications after PGI surgery.

A random-effects meta-analysis with restricted maximum likelihood was used to summarise included literature. Adult and paediatric populations were analysed separately. Mean difference (MD) with 95% confidence intervals was performed. When standard deviations were not available, these were estimated using the Walter and Yao method or the Method for Unknown Non-Normal Distributions (MLN) depending on available summary statistics (median, range or interquartile range).^10,11^ Heterogeneity between studies was quantified using the modified Higgin’s 𝐼^2^ statistic (𝐼_A_^2^).^12^

When considering the true effect size in a meta-analysis, as the sampling distribution of the observed effect is dependent on sample size, this converges to the true effect size as sample size increases. Hence, regardless of the size of the underlying heterogeneity between studies, the widely used Higgins 𝐼^2^ statistic will approximate 100% as sample size increases.^13^ We used the 𝐼_A_^2^statistic as a measure for quantifying heterogeneity in our meta-analysis as it is an unbiased estimator that also has the property of sample size invariance distinguishing it from the traditional Higgins 𝐼^2^statistic.^12^

We conducted subgroup analyses based on application of MMC and geographical location of the studies. A p-value less than 0.05 was considered statistically significant. All analyses were performed using R Statistical Software (v4.2.2; R Core Team 2021).

## Results

### Study selection

A total of 131 articles were retrieved following systematic search of the databases. After removing duplicates and assessing for eligibility, 24 articles were included in this meta-analysis, four articles for children and 20 articles for adults. One article (Hu et al. 2024) was published in Chinese.^14^ Two articles were excluded as the data were published in subsequent follow-up studies.^15,16^ The literature search is summarised in a PRISMA flow chart (Figure 1).

**Figure 1.**
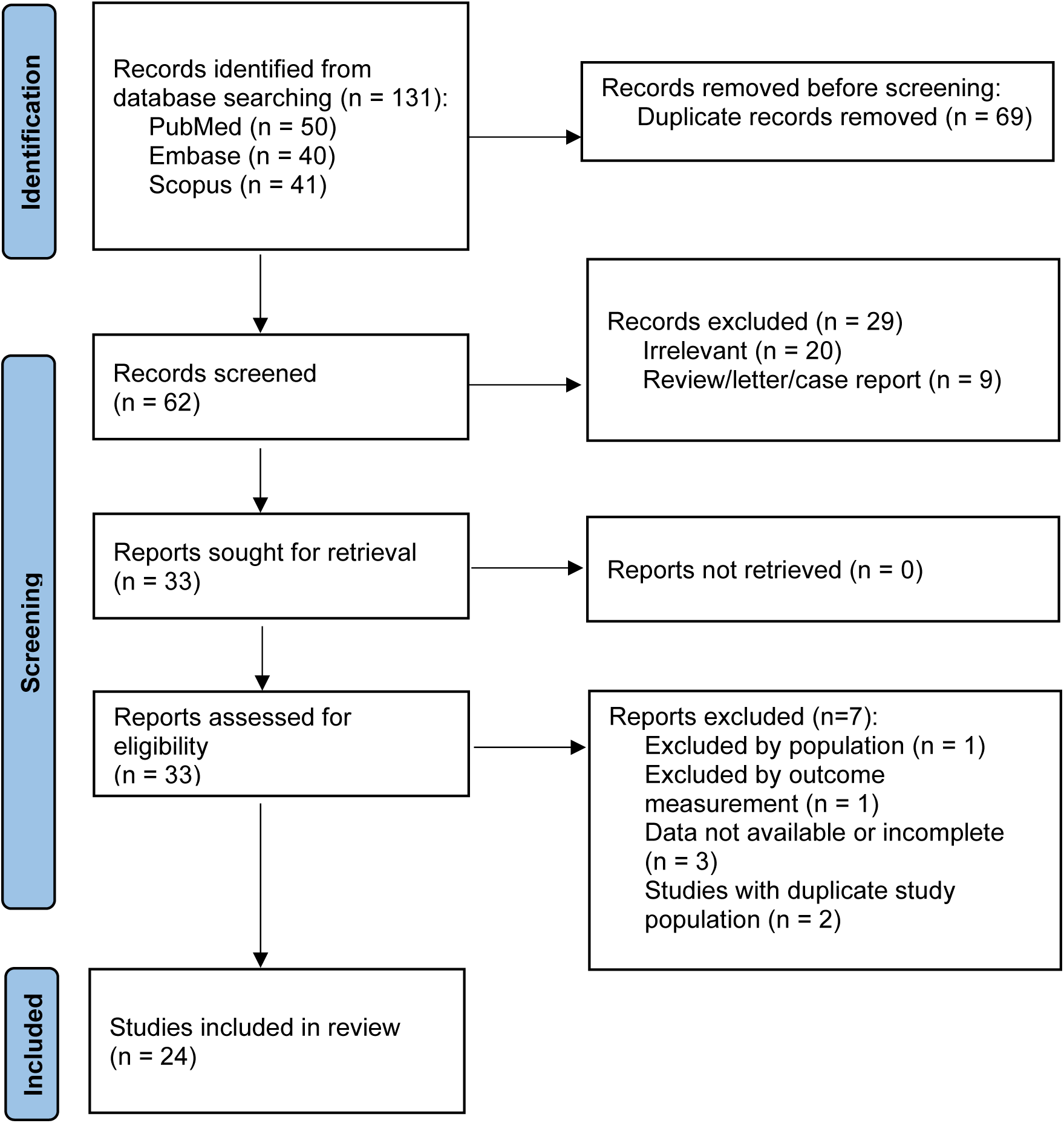
PRISMA flow chart.

### Study demographics

Table 1 summarises the characteristics of the included studies.

**Table 1.**
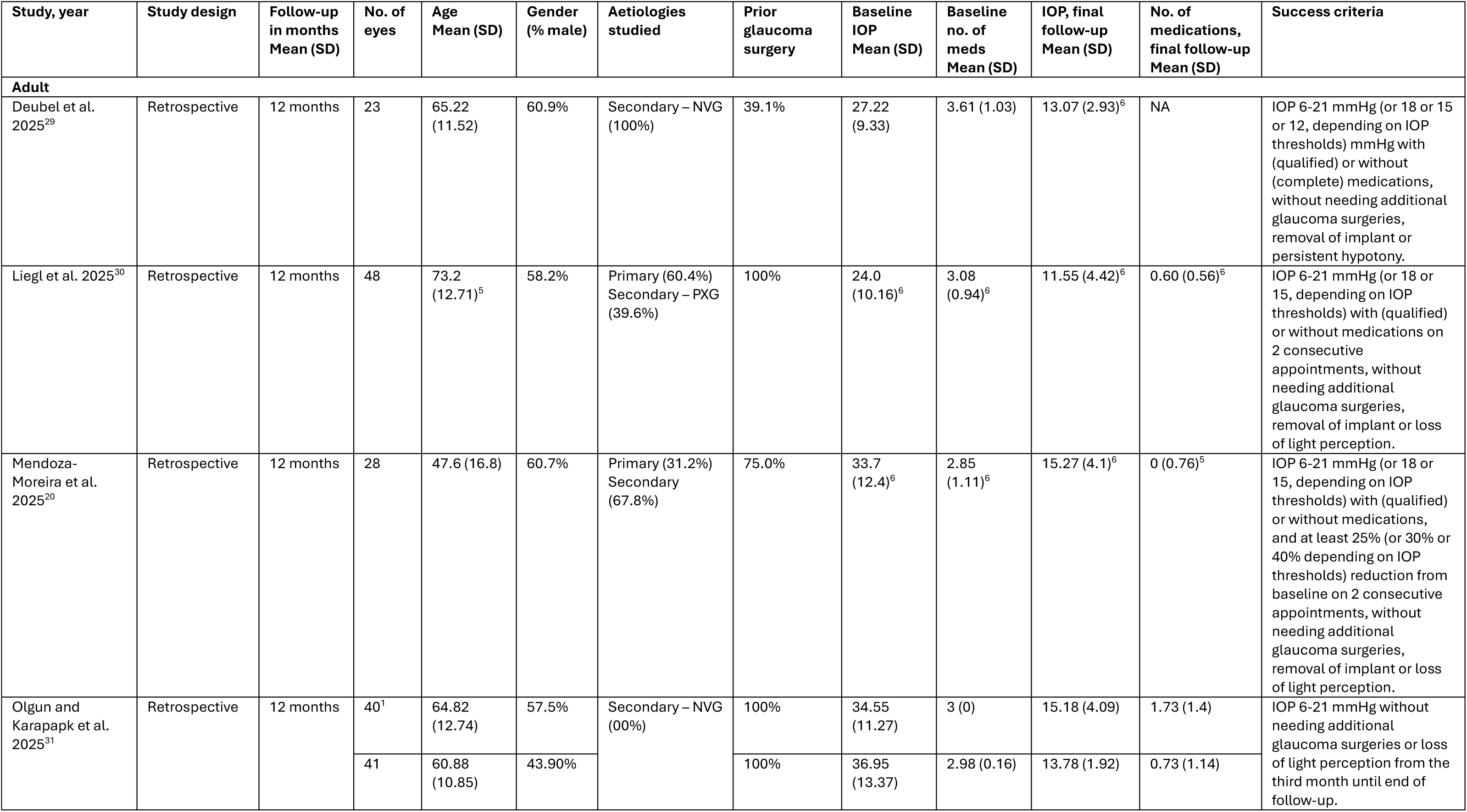
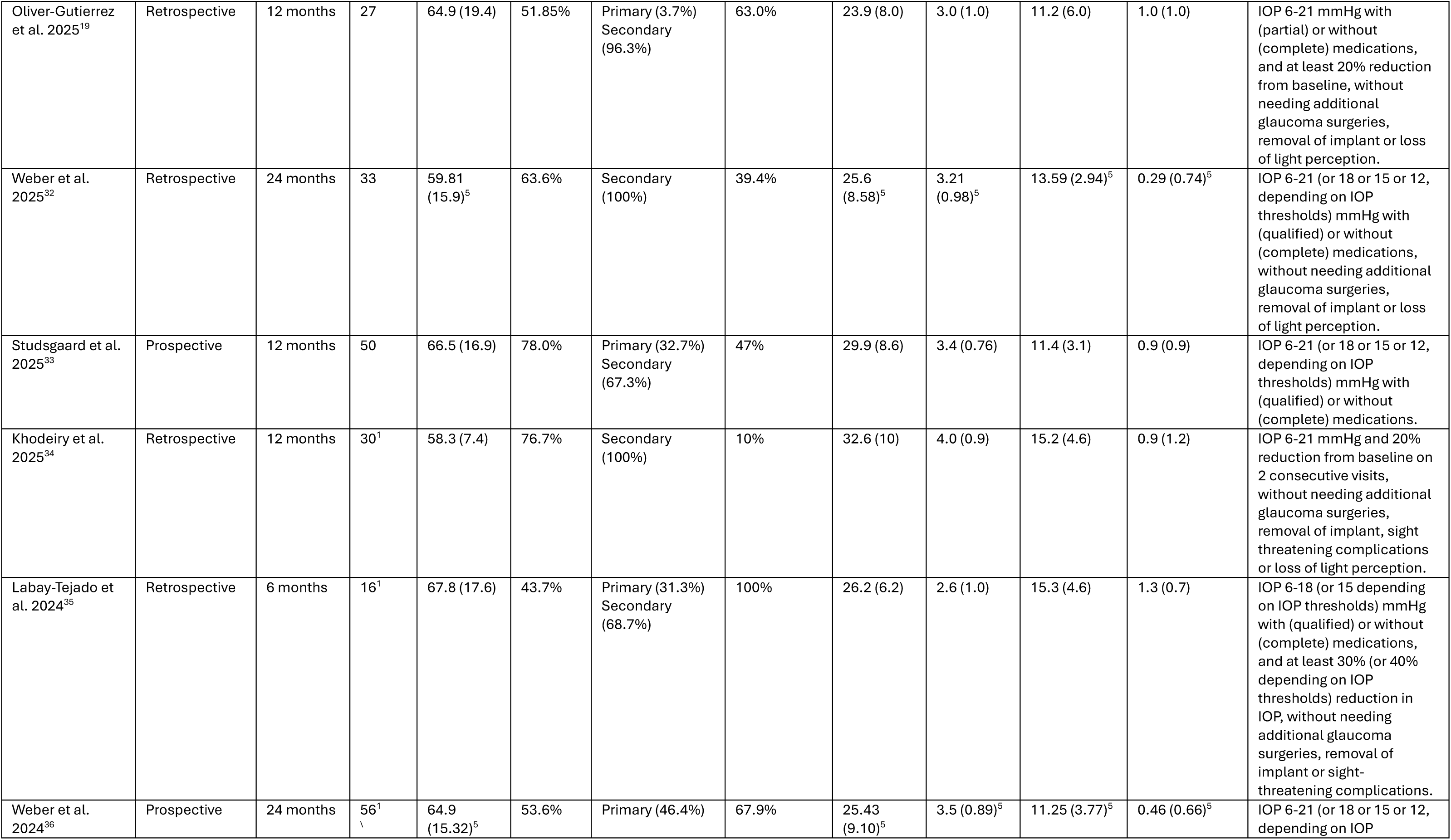
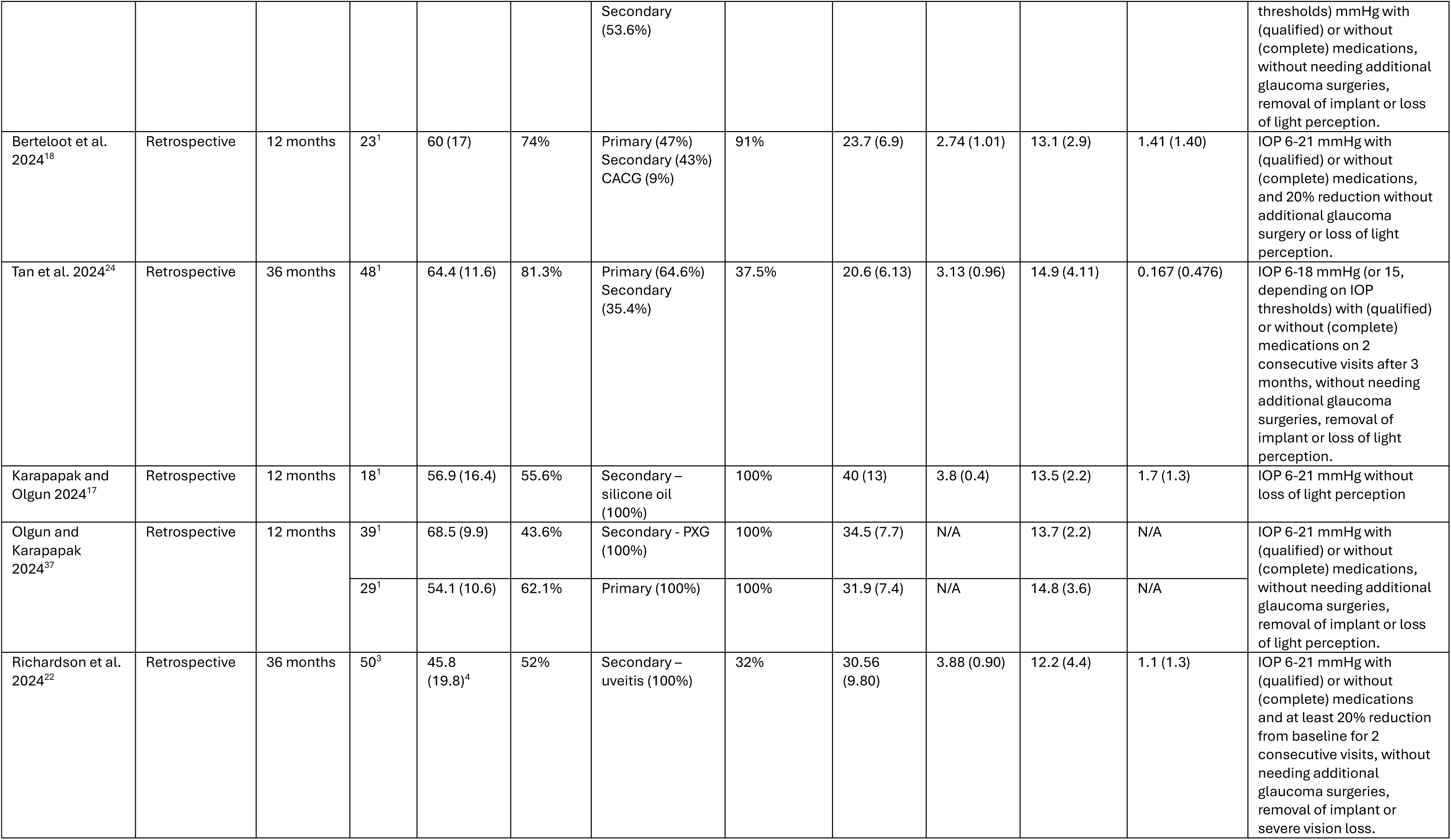
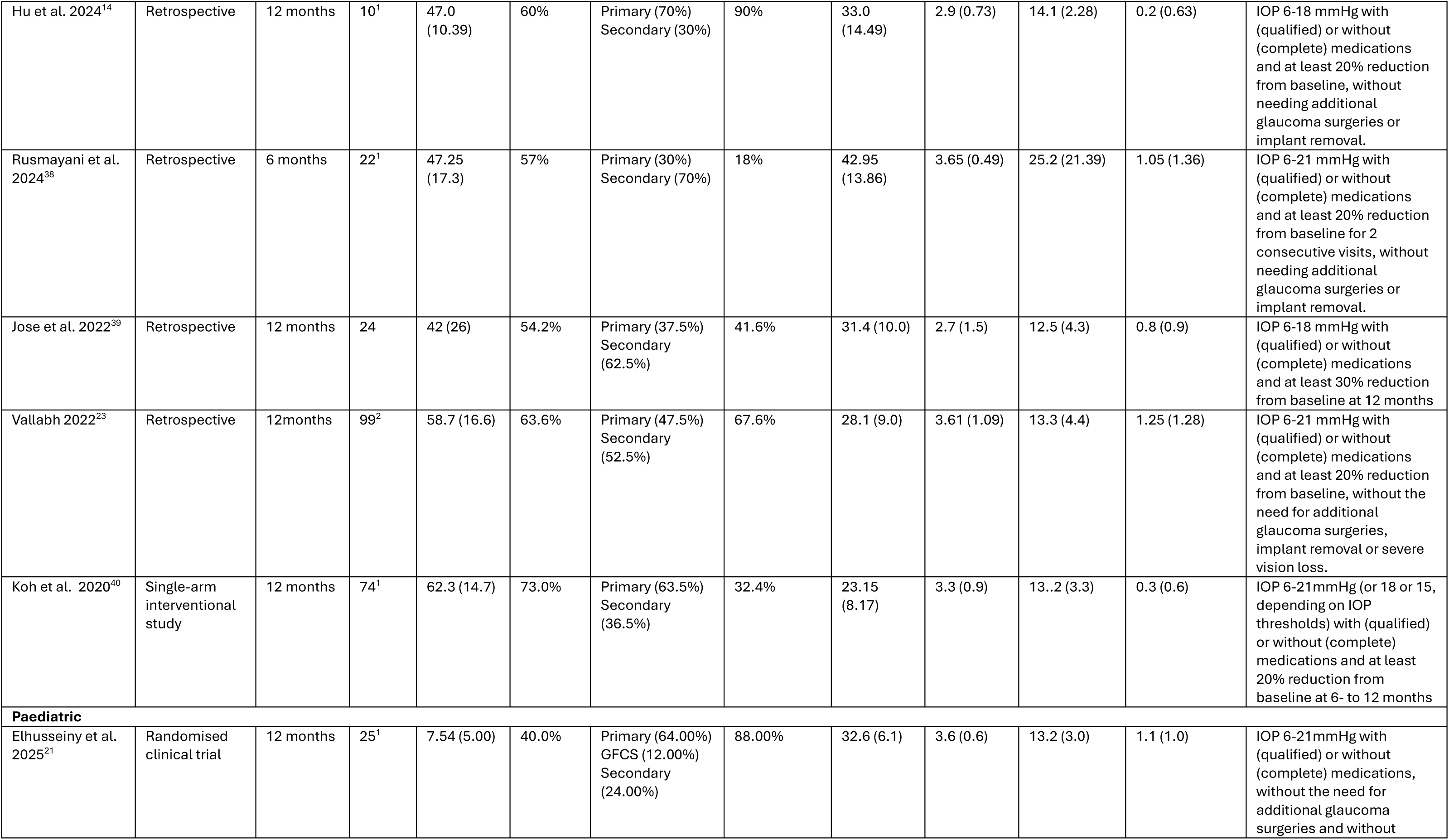
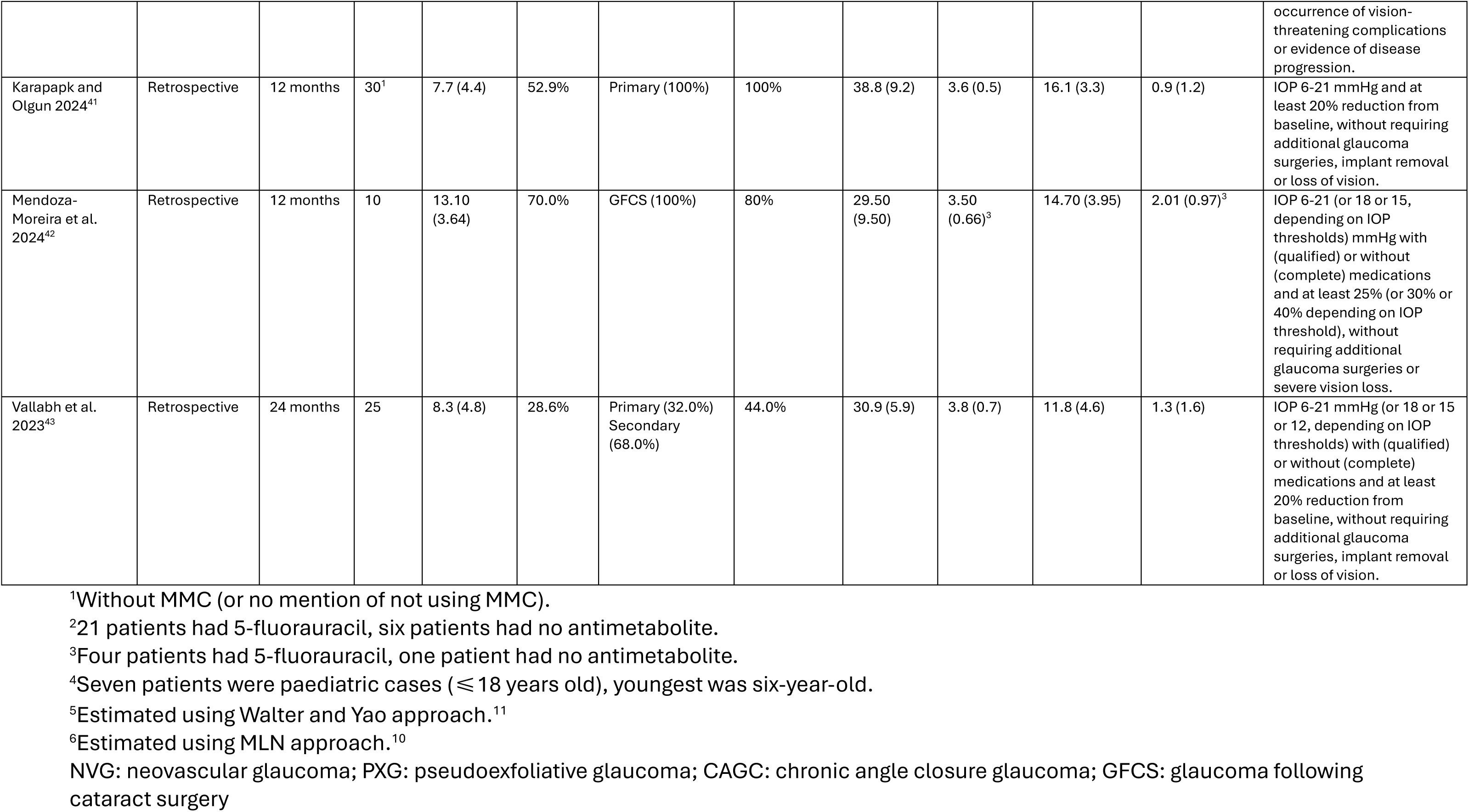
Baseline characteristics of included studies.

In total, 836 eyes were included (90 children). Five of the included studies compared PGI with other glaucoma drainage devices (Ahmed Glaucoma Valve and Baerveldt Glaucoma Implant).^17–21^ Follow-up period ranges from 6 to 48 months. Twenty-one studies had 12- month follow-up, and only one study has a 48-month follow-up.

The use of Mitomycin C (MMC) was dependent on the studies, with 458 of 836 eyes receiving it as part of the PGI surgical protocol, with concentrations between 0.2 to 0.5 mg/mL, placed for between 90 seconds to 3 minutes.

For the adult population, primary open angle glaucoma (POAG) was a common indication for PGI accounting for between 13 and 60% of participants in the included studies. Secondary glaucoma included pseudoexfoliation glaucoma (PXG), neovascular glaucoma (NVG), uveitic glaucoma and glaucoma post-vitreoretinal surgery. Most patients, 533 of 836 eyes (63.6%) had previous glaucoma surgery. Mean age ranges from 42 to 73 years. Mean baseline IOP was 28.96 ± 2.23 mmHg. The mean number of IOP-lowering medications used was 3.31 ± 0.24.

For the paediatric population, primary congenital glaucoma was the main indication for PGI in three of the included study. Mendoza-Moreira et al 2024 only included children with glaucoma following cataract surgery (GFCS). Of the four included studies, 71 of 90 eyes (78.9%) had previous glaucoma procedure. Mean age ranges from 7.5 to 13.1 years old. Mean baseline IOP was 33.85 ± 4.01 mmHg. The mean number of pre-operative IOP-lowering medications was 3.64 ± 0.31.

### Intraocular pressure

A total of 24 studies were included in the meta-analysis. For both adults and children, there was a significant reduction in IOP 1 week after PGI surgery, with the effect maintained at 12 months follow-up. Figures 2 and 3 summarise these findings.

**Figure 2.**
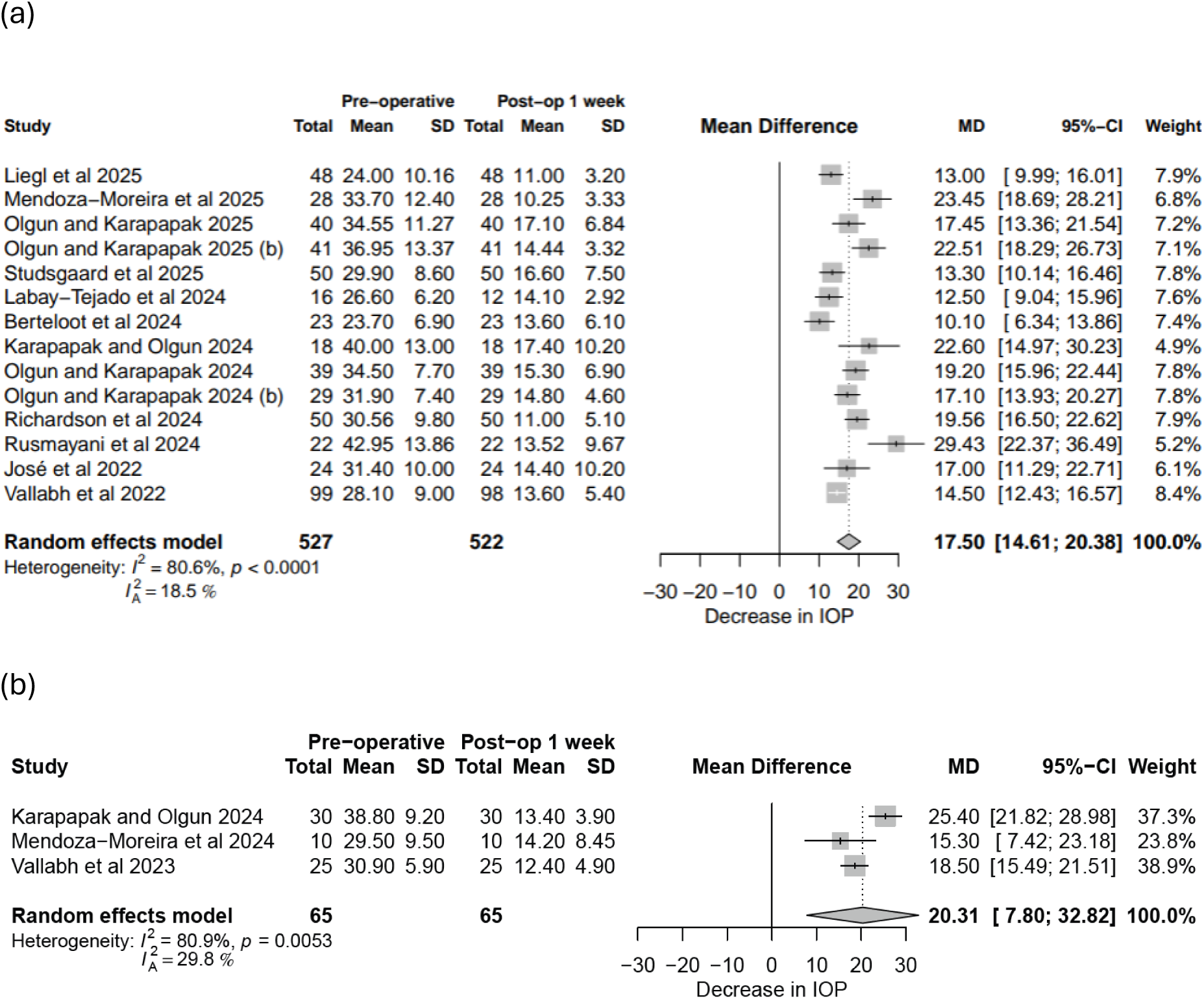
Forest plot comparing pre- and post-operative (1 week) IOP with PGI: (a) Adults, (b) Children.

**Figure 3.**
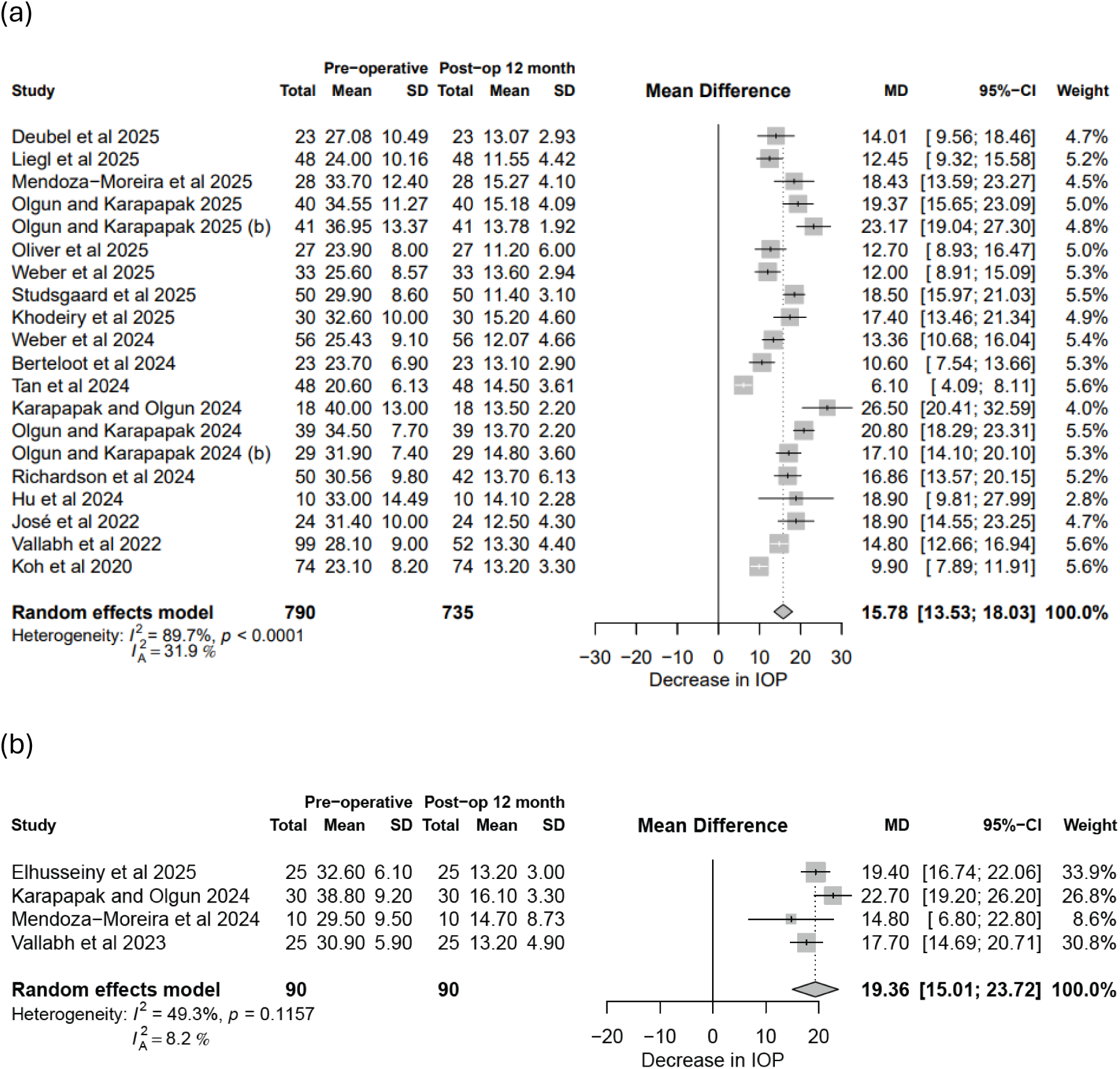
Forest plot comparing pre- and post-operative (12 months) IOP with PGI: (a) Adults, (b) Children.

A total of 535 adult and 65 paediatric eyes were included, with only 1 adult loss to follow-up at 1 week. The IOP was significantly lower after PGI surgery with a mean difference of 17.48mmHg (95% CI: 14.59 – 20.37) and 20.31mmHg (95% CI: 7.80 – 32.82) in adults and children respectively. The modified heterogeneity test showed that there was mild heterogeneity after accounting for sample size, with 𝐼_A_^2^ < 30%.

At 12 months post-operative, the IOP reduction remained statistically significant for both adults and children (Figure 3). In total 790 adult eyes underwent preoperative IOP testing and 735 had IOP recorded at 12 months. All 90 paediatric patient remained in the study by 12 months follow-up. There was again a significant reduction of IOP with mean difference of 15.78mmHg (95% CI: 13.53, 18.03) and 19.36mmhg (95% CI: 15.01, 23.72) in adults and children respectively.

Figure 4 shows the trend in mean IOP reduction over follow-up duration. There were 3 and 4 studies describing changes in IOP 18- and 24-months after initial PGI surgery respectively, resulting in wider confidence intervals. Generally, for both adults and paediatric patients, we can see an immediate reduction in mean IOP from day 1 post-op, which remained relatively stable through the follow-up period after 3-months.

**Figure 4.**
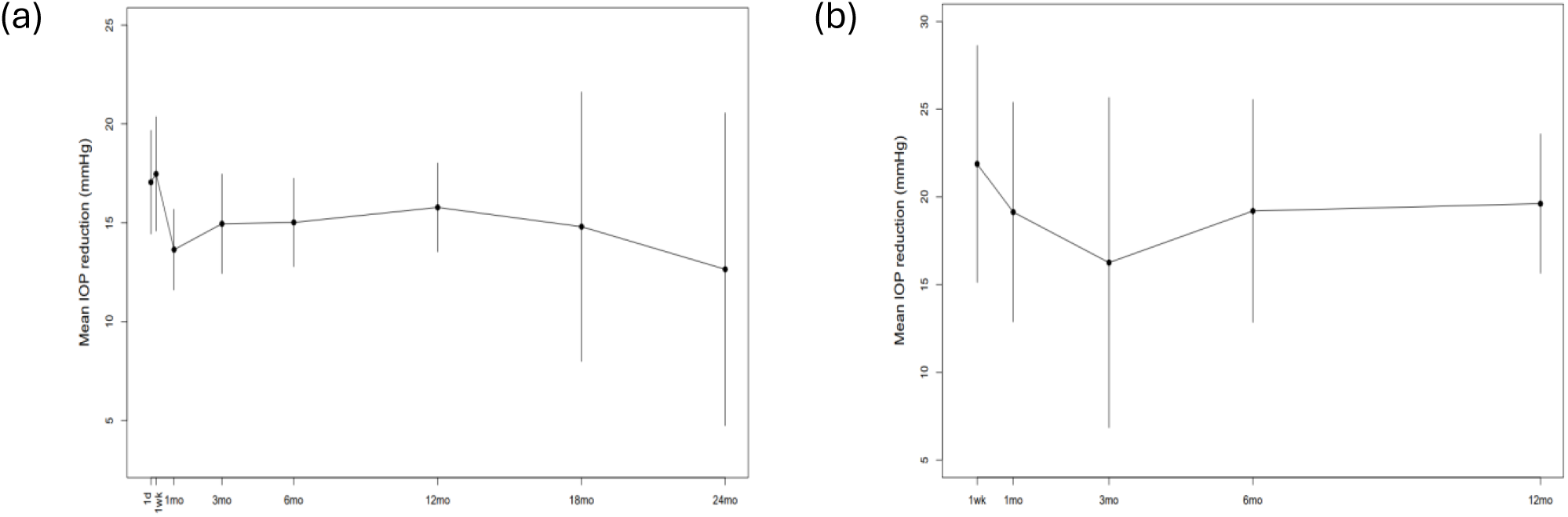
Mean IOP reduction over follow-up duration: (a) Adults, (b) Children.

### Glaucoma medications

The average reduction in number of postoperative glaucoma medications after PGI implantation was 2.37 at 1 month (95% CI: 2.00 – 2.74) and 2.56 at 12 months (95% CI: 2.28 – 2.83) for adults. For the paediatric population, the mean reduction in number of medications at 1 month and 12 months were 3.17 (95%CI: -2.55 – 8.88) and 2.60 (95% CI: 2.13 – 3.07). Figures 5 and 6 summarise these findings.

**Figure 5.**
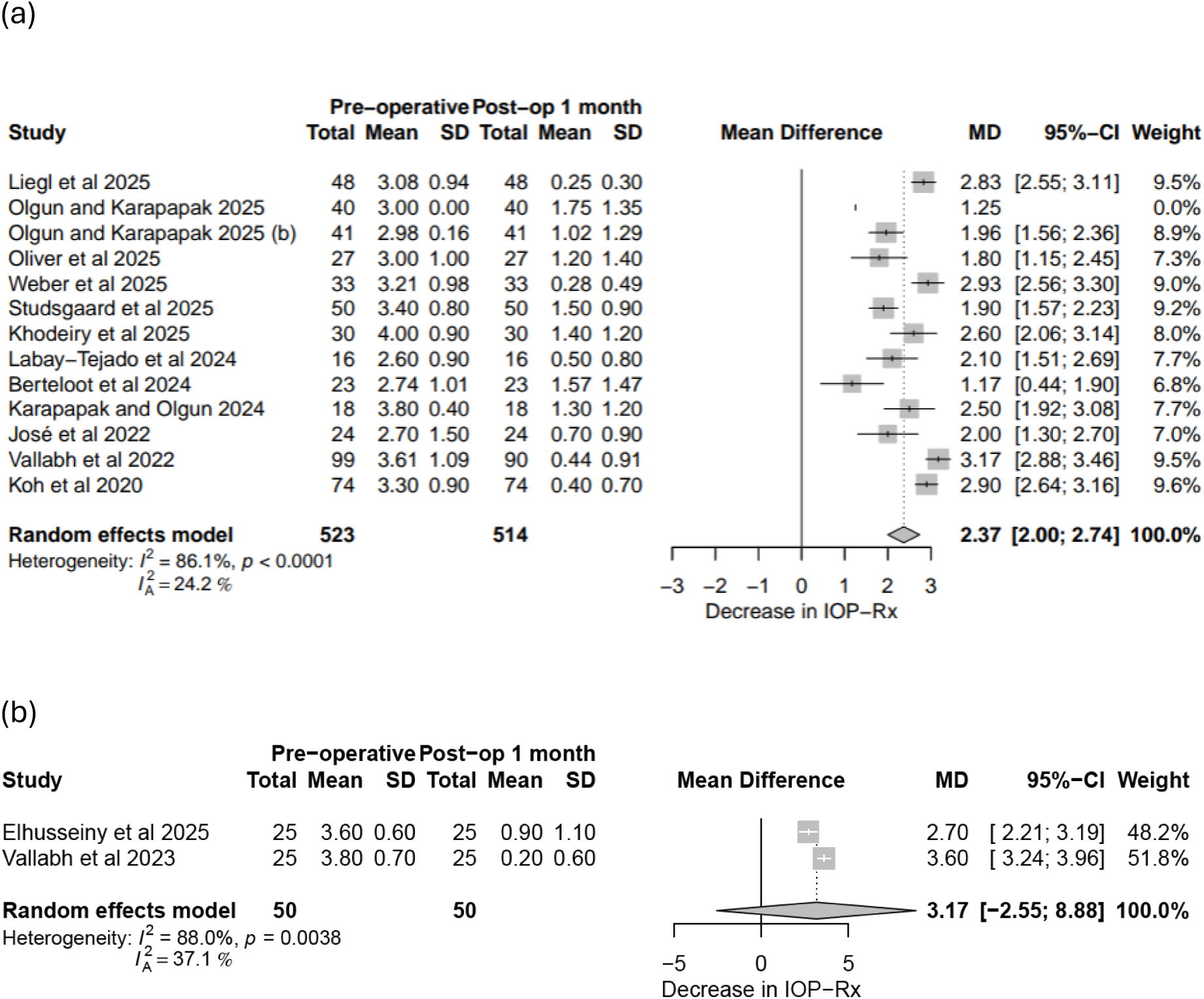
Forest plot comparing pre- and post-operative (1 month) number of anti-glaucoma medications after PGI: (a) Adults, (b) Children.

**Figure 6.**
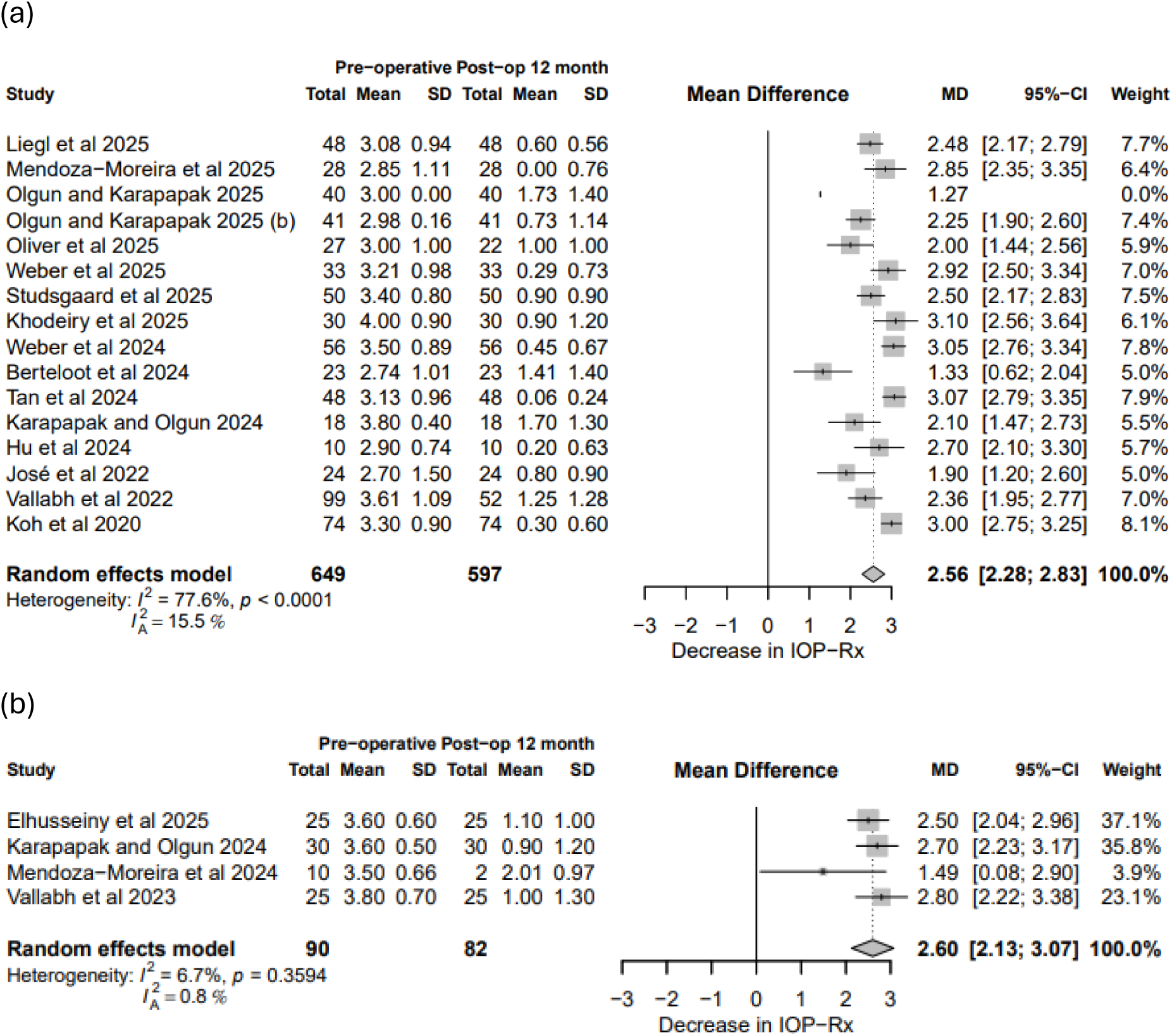
Forest plot comparing pre- and post-operative (12 months) number of anti-glaucoma medications after PGI: (a) Adults, (b) Children.

Mendoza-Moreira et al. had 80% of patients (n=8) lost to follow-up at the 12 months mark. We have included this in our meta-analysis, but note that the observed effect size differs from the other three studies, and contributed 6.7% to the estimated heterogeneity.

Figure 7 shows the trend in mean reduction in anti-glaucoma medications over follow-up duration. The associated wide confidence intervals with some time-points is due to the small number of studies included. For adults, the mean reduction in anti-glaucoma medications appear to increase over follow-up duration until 18-months after initial surgery before plateauing off. For paediatric population, the mean reduction in number of anti-glaucoma medications remained relatively stable from 1-month post-operative.

**Figure 7.**
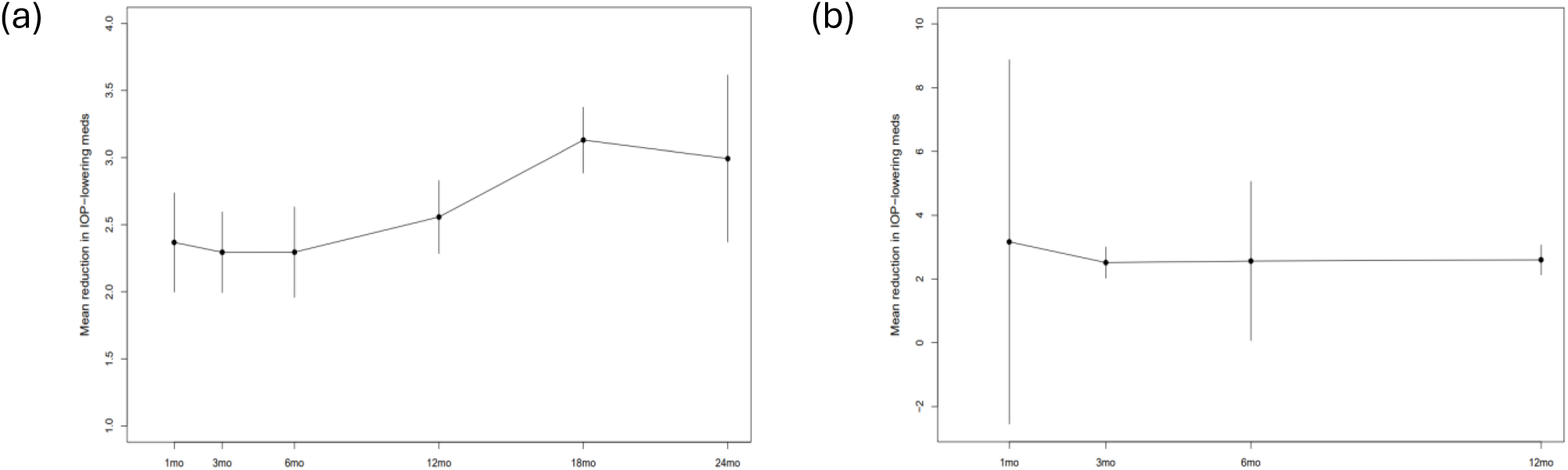
Mean reduction in anti-glaucoma medications over follow-up duration: (a) Adults, (b) Children.

### Mitomycin-C (MMC)/anti-metabolites use

A total of 10 studies used MMC (or 5-fluorauracil (5-FU)) during PGI surgery. Richardson et al. 2024 and Vallabh et al. 2022 had 10% and 27% of patients not receiving MMC due to supply issue, with most of these patients receiving 5-FU instead.^22,23^ The concentration of MMC ranges between 0.2 to 0.5 mg/mL, applied for between 90 seconds to 3 minutes. At 12-months follow-up, for both adult and paediatric patients, there were no significant difference in IOP reduction between those who had MMC as part of the procedure and those who did not (Figure 8). There were also no significant differences in reduction of number of anti-glaucoma medications at 12-months follow-up (Adult: 𝜒^2^ = 0.00, 𝑝 = 0.9587; Paediatric: 𝜒^2^ = 0.20, 𝑝 = 0.6512).

**Figure 8.**
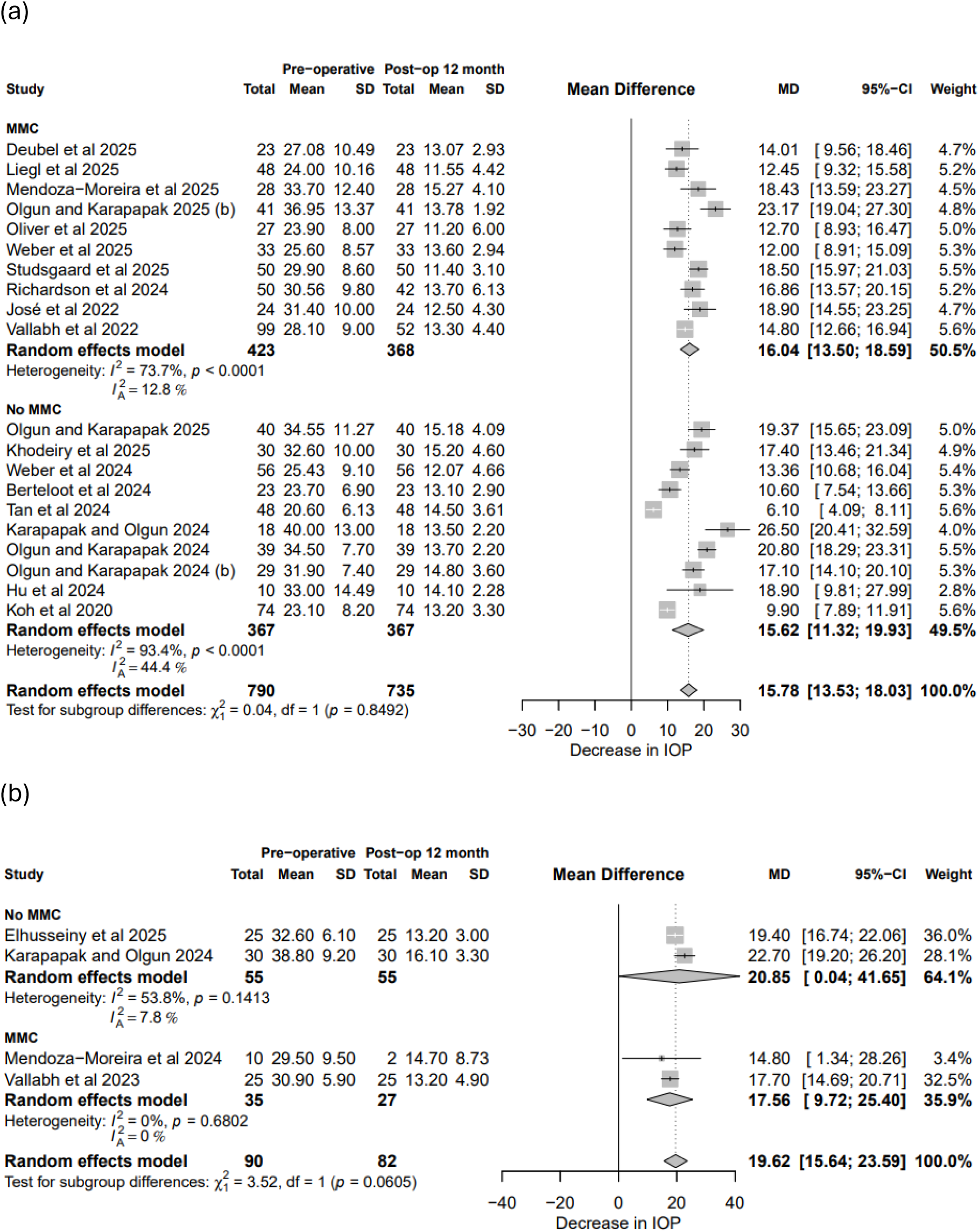
Forest plot comparing pre- and post-operative (12 months) IOP after PGI, subgroup by use of MMC during procedure: (a) Adults, (b) Children.

**Figure 9.**
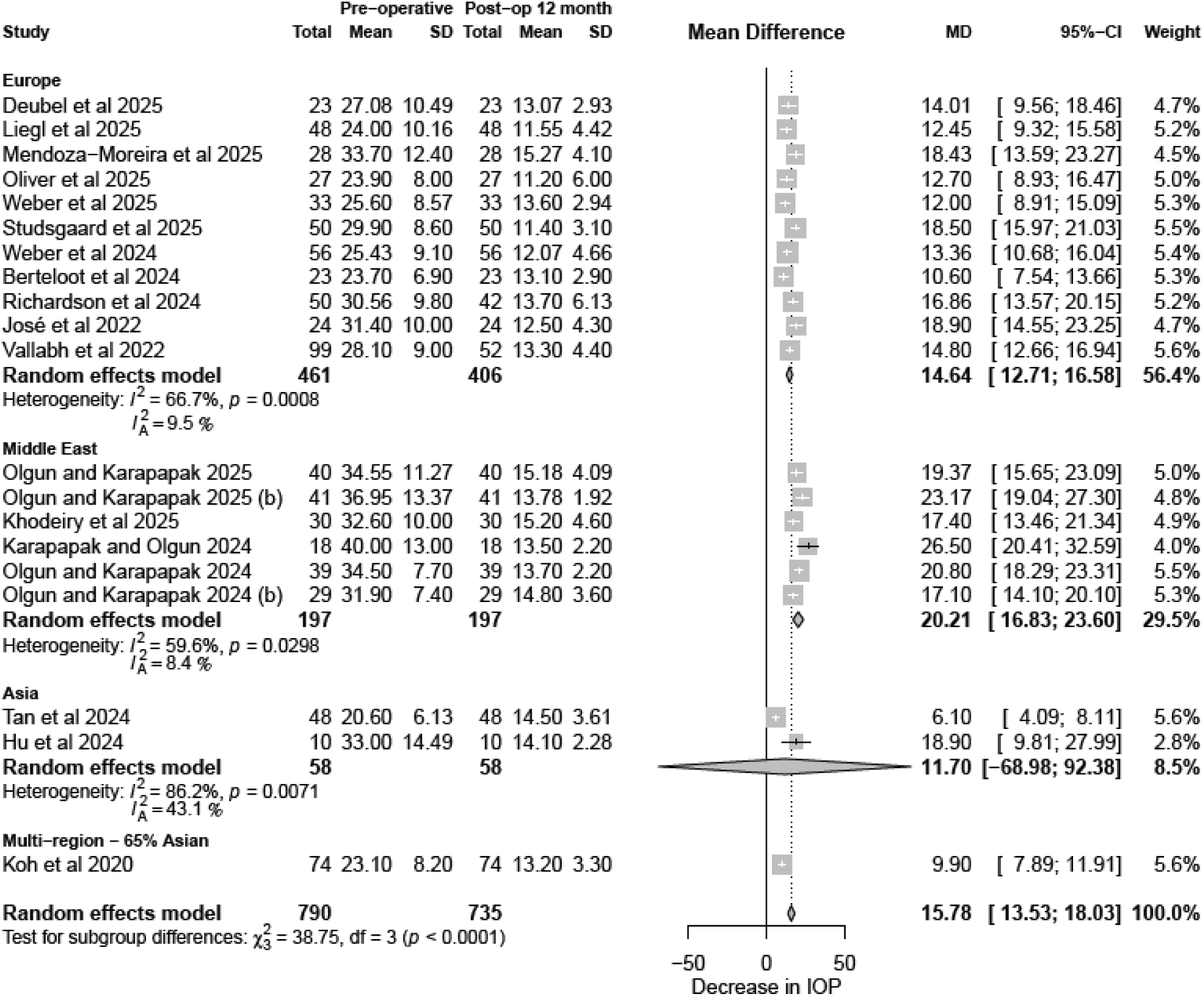
Forest plot comparing pre- and post-operative (12 months) IOP after PGI for adults, subgroup by region.

### Region

A total of 18 studies investigating the adult population were included in this subgroup analysis by region. Eleven studies were from Europe, four studies were from Middle East, two studies were from Asia, and one was an international multi-centre study with participants from Singapore, Thailand, Hong Kong, UK and Malaysia. The multi-centre study by Koh et al. 2020 had 65% of the participants identifying as Asian ethnicity.

There was significant difference in mean reduction of IOP at 12-months follow-up between population from different geographic region. Study populations located in Middle East had higher mean IOP reduction compared to other regions. Hu et al. 2024 only included 10 participants in their observational cohort study, all of whom had much higher pre-operative IOP readings compared to other studies with predominantly Asian participants.^7,24^ The resulting wide confidence interval showed no significant reduction in mean IOP at 12-months follow-up (Mean difference 11.70 mmHg; 95% CI: -68.98 – 92.38). Figure 8 summarises these findings. Sensitivity analysis with this study removed from the analysis strengthen the conclusion that there were differences in mean IOP reduction at 12-months by geographic location (𝜒^2^ = 85.19 vs 𝜒^2^ = 38.75; df = 3).

There was also a significant difference in mean reduction of the number of anti-glaucoma medications at 12-months follow-up between regions (𝜒^2^ = 9.21, 𝑑𝑓 = 3, 𝑝 = 0.0266). Patients from Asia had larger reduction in the number of anti-glaucoma medications at 12-months after initial PGI surgery, compared to other population.

### Complications

Complications are reported as early (≤ 3 months) or late (> 3 months). In the adult population, hyphema (9.81%), hypotony defined as IOP < 6mmHg (9.01%) and flat or shallow anterior chamber without numerical hypotony (8.57%) were the most common early adverse events after PGI surgery. In terms of complications associated with hypotony, choroidal detachment occurred was reported at 5.81%, maculopathy at 1.37% and shallow anterior chamber at 3.25%. Most cases of hypotony are transient and resolve without needing intervention. 3% of patients developed diplopia in the early post- operative period, with only 1 patient eventually receiving surgical intervention. Tube/stent exposure was reported in 5.35% of patients from 8 studies, many of which required intervention. Tube occlusion (4.71%) can be due to viscoelastic, fibrin, or blood.

Late complications tend to be more serious. Delayed tube/stent exposure was seen in 7.28% of cases, requiring revision surgery or explantation. Corneal decompensation was observed in 4.49% of cases. Endophthalmitis was only reported in one subject. Table 2 summarises the reported complications in adults.

**Table 2:**
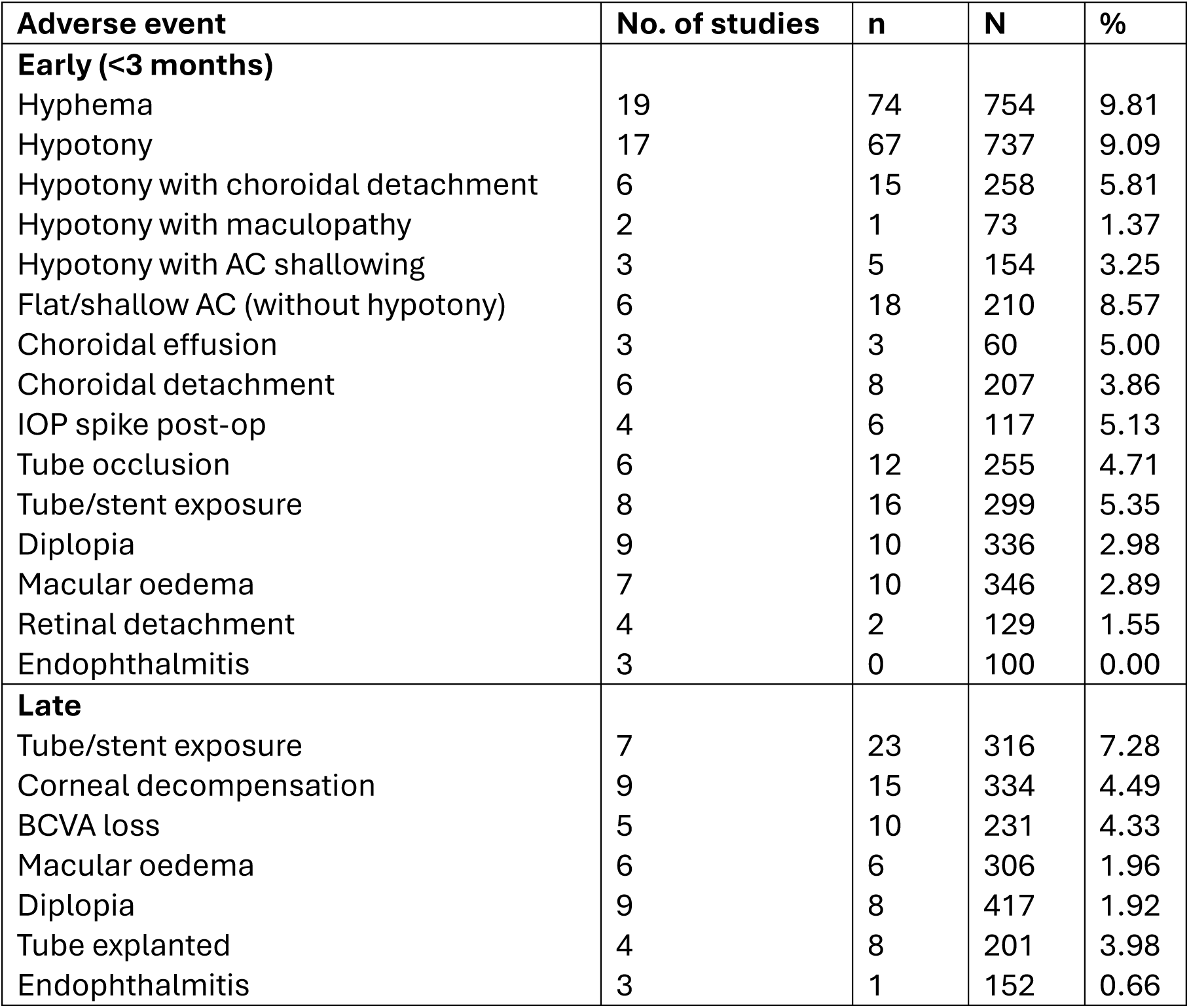
Adverse events – adults.

| **Adverse event** | **No. of studies** | **n** | **N** | **%** |
| --- | --- | --- | --- | --- |
| **Early (<3 months)** |  |  |  |  |
| Hyphema | 19 | 74 | 754 | 9.81 |
| Hypotony | 17 | 67 | 737 | 9.09 |
| Hypotony with choroidal detachment | 6 | 15 | 258 | 5.81 |
| Hypotony with maculopathy | 2 | 1 | 73 | 1.37 |
| Hypotony with AC shallowing | 3 | 5 | 154 | 3.25 |
| Flat/shallow AC (without hypotony) | 6 | 18 | 210 | 8.57 |
| Choroidal effusion | 3 | 3 | 60 | 5.00 |
| Choroidal detachment | 6 | 8 | 207 | 3.86 |
| IOP spike post-op | 4 | 6 | 117 | 5.13 |
| Tube occlusion | 6 | 12 | 255 | 4.71 |
| Tube/stent exposure | 8 | 16 | 299 | 5.35 |
| Diplopia | 9 | 10 | 336 | 2.98 |
| Macular oedema | 7 | 10 | 346 | 2.89 |
| Retinal detachment | 4 | 2 | 129 | 1.55 |
| Endophthalmitis | 3 | 0 | 100 | 0.00 |
| **Late** |  |  |  |  |
| Tube/stent exposure | 7 | 23 | 316 | 7.28 |
| Corneal decompensation | 9 | 15 | 334 | 4.49 |
| BCVA loss | 5 | 10 | 231 | 4.33 |
| Macular oedema | 6 | 6 | 306 | 1.96 |
| Diplopia | 9 | 8 | 417 | 1.92 |
| Tube explanted | 4 | 8 | 201 | 3.98 |
| Endophthalmitis | 3 | 1 | 152 | 0.66 |

In the paediatric population, the most reported early complication was hypotony (6.7%, 4/60 eyes). Among paediatric patients with refractory primary congenital glaucoma, Karapapak and Olgun reported 10% (3/30 eyes) having choroidal detachment in the early postoperative period. Tube exposure, re-stenting, revision and explantation were between 3 – 4%. There were no reported cases of endophthalmitis, diplopia, strabismus or cataract complications.

### Risk of bias assessment

Results from bias assessment are found in Supplementary 3. Most of the included observational studies are single cohort study and retrospective. Strict inclusion criteria of having complete follow-up, i.e. excluding patients who did not complete follow-up was considered as serious risk of selection bias. Risk of measurement biases were low given standardised methods of IOP measurements. Misclassification of intervention is unlikely given the nature of the intervention (PGI surgery). All included studies were judged as low to moderate risk.

### Comparison with other glaucoma drainage device

In the adult population, two studies compared PGI with Ahmed glaucoma valve (AGV) and Baerveldt glaucoma implant (BGI) respectively. At one day postoperative, the mean IOP reduction was 64% and 61% for PGI and AGV respectively.^17,20^ At one week after surgery, mean IOP reduction was 65% and 57% for PGI and AGV respectively. Berteloot et al. reported 23% mean IOP reduction for patients with BGI compared to 43% for those who had PGI.^18^ At 12 months follow-up, the mean IOP reduction was 61% and 53% for PGI and AGV respectively.^17,20^ For studies comparing BGI and PGI, the mean IOP reduction was 48% and 59% for PGI and BGI respectively.^18,19^

Among paediatric patients, there was only one study comparing PGI with AGV. The study reported significantly higher mean IOP at 1 month postoperative for PGI (14.6 ±6.2 mmHg) compared to AGV (9.05 ±5.6 mmHg), representing a 70% and 55% reduction in mean IOP respectively, but no statistically significant differences at 3, 6 and 12 months.^21^

## Discussion

This systematic review and meta-analysis of 24 studies found that PGI is associated with early reduction in mean IOP with the effect sustained up to 36-months for adults, and 12- months for children. At one week after initial PGI surgery, the mean IOP is reduced by 17.48mmHg (95% CI: 14.59, 20.37) and 20.31mmHg (95% CI: 7.80, 32.82) for adults and children, respectively. This represents 55.5% and 60.6% reduction from baseline values. Mean reduction in the number of anti-glaucoma medications at 1 month after PGI surgery was sustained by 12-months follow-up. At 12-months follow-up, the average reduction in the number of anti-glaucoma medications were 79.2% in adults and 71.4% in children, compared to pre-surgery. Application of MMC during the surgical procedure did not significantly alter mean IOP reduction. There is significant association between mean IOP reduction and geographic location, with greater reductions for studies conducted in Middle East.

Two of the most used glaucoma drainage devices are the AGV and BGI. The Ahmed Versus Baerveldt (AVB) study demonstrated a significantly lower mean IOP at the 1-day, 1-week and 2-week follow-up visits for patients with AGV compared to BGI. However, at the 1- year follow-up, BGI showed a greater IOP-lowering effect.^25^ These findings are consistent with those from the Ahmed Baerveldt Comparison (ABC) study, which also reported significantly lower mean IOP at the 1-day and 1-week follow-up visits for AGV, with BGI achieving a greater reduction in IOP at the 1-month follow-up. At 12-months, pooled data from the ABC and AVB studies showed mean IOPs of 15.9 ± 5.3mmHg and 13.6 ± 5.9mmHg for AGV and BGI respectively, representing a 50% reduction in mean IOP from baseline.^26^ Results from this meta-analysis showed comparable mean IOP reductions, with a 55% reduction at 1-week, 58% at 12-months, and 46% at 36-months follow-up for adults. In children, mean IOP reduction was 63% at 1-week, 58% at both 1-month and 12- months follow-up.

The BGI has been shown to be associated with significantly lower number of anti-glaucoma medications at the final follow-up compared with AGV.^26,27^ Pooled results from the ABC and ABV studies demonstrated a 64% reduction in the mean number of glaucoma medications at 6-months for BGI, compare with a 48% reduction for AGV. Findings from our meta-analysis indicated greater reductions in number of glaucoma medications with the PGI, with mean reductions of 74% at 1-month and 71% at 6-months in adults, and 85% at 1-month and 70% at 6-moths in children. At 12-months, mean reductions in glaucoma medications for PGI were 79% in adults and 71% in children, compared with 58% for BGI and 45% for AGV. The PGI demonstrated a sustained reduction in medication use through 36 months, with mean reductions of 88% in adults. This compares favourably with corresponding mean reductions of 61% and 42% for BGI and AGV respectively.^26^

Adverse events were included in the analysis only when explicitly reported in the primary studies, to minimise risk of underestimation. The rates of adverse events associated with the PGI are comparable to that of other glaucoma drainage device. The incidence of hypotony maculopathy was 1.37% for PGI, lower than the 5.8% reported for BGI.^28^ Compared with other non-randomised studies, choroidal effusion was reported in 5% of eyes following PGI implantation, compared with reported rates of 4.3 – 12.0% for AGV and 8.2 – 10.3% for BGI.^27,28^ Vision loss, defined as a reduction of more than two lines on the Snellen chart, was observed in 6.6% (8/122) of eyes with PGI implantation, which is lower than rates reported for AGV (13.6%) and BGI (23.1%).^28^ Risk of diplopia is also lower with PGI at 2.98% (10/336 eyes) in the early and 1.92% (8/417 eyes) in the late postoperative period, compared to pooled estimates of 15.4% for AGV and 5.9% for BGI.^28^ Rate of endophthalmitis following PGI implantation was similarly lower than that reported for AGV (0.95%) and BGI (0.63%).^27^

This study represents the first meta-analysis of outcomes associated with the PGI. However, several limitations must be acknowledged. With the exception of Elhusseiny et. al. (2025), all included studies were retrospective in nature. The number of eyes with follow-up beyond 12 months was relatively small, limiting the strength of conclusions drawn regarding long-term outcomes. There is also considerable heterogeneity across studies. MMC concentrations were not standardised across studies. Race has long been recognised as a significant risk factor for glaucoma and its progression; however, we attempted to account for this source of heterogeneity through subgroup analysis by geographic region. The patient population included in this meta-analysis have high prevalence of secondary glaucoma, with many patients having had previous glaucoma surgeries, which may influence generalisability of the results especially among patients being considered for de novo glaucoma surgeries.

In conclusion, the Paul glaucoma implant offers an effective and safe surgical option for glaucoma management in both adult and paediatric populations. The mean IOP reduction was comparable to the Ahmed glaucoma valve and Baerveldt glaucoma implant, while needing much fewer medications across all follow-up periods. Incidence of post-operative adverse events were generally lower compared to other glaucoma drainage devices. However, further larger, longer-term prospective studies with direct comparisons between PGI and other glaucoma drainage devices are needed to assess long-term efficacies and complications. Based on current evidence, the PGI should be considered as an alternative to other glaucoma drainage devices. Ultimately, the selection of a suitable device is multifactorial – taking into consideration patient’s risk factors, individual IOP target, urgency of achieving IOP target, compliance with glaucoma medications, and also surgeons’ experience with each device.

## Data Availability

All data produced in the present work are from published studies and are available upon reasonable request to the authors.

---

**Supplementary 1:**
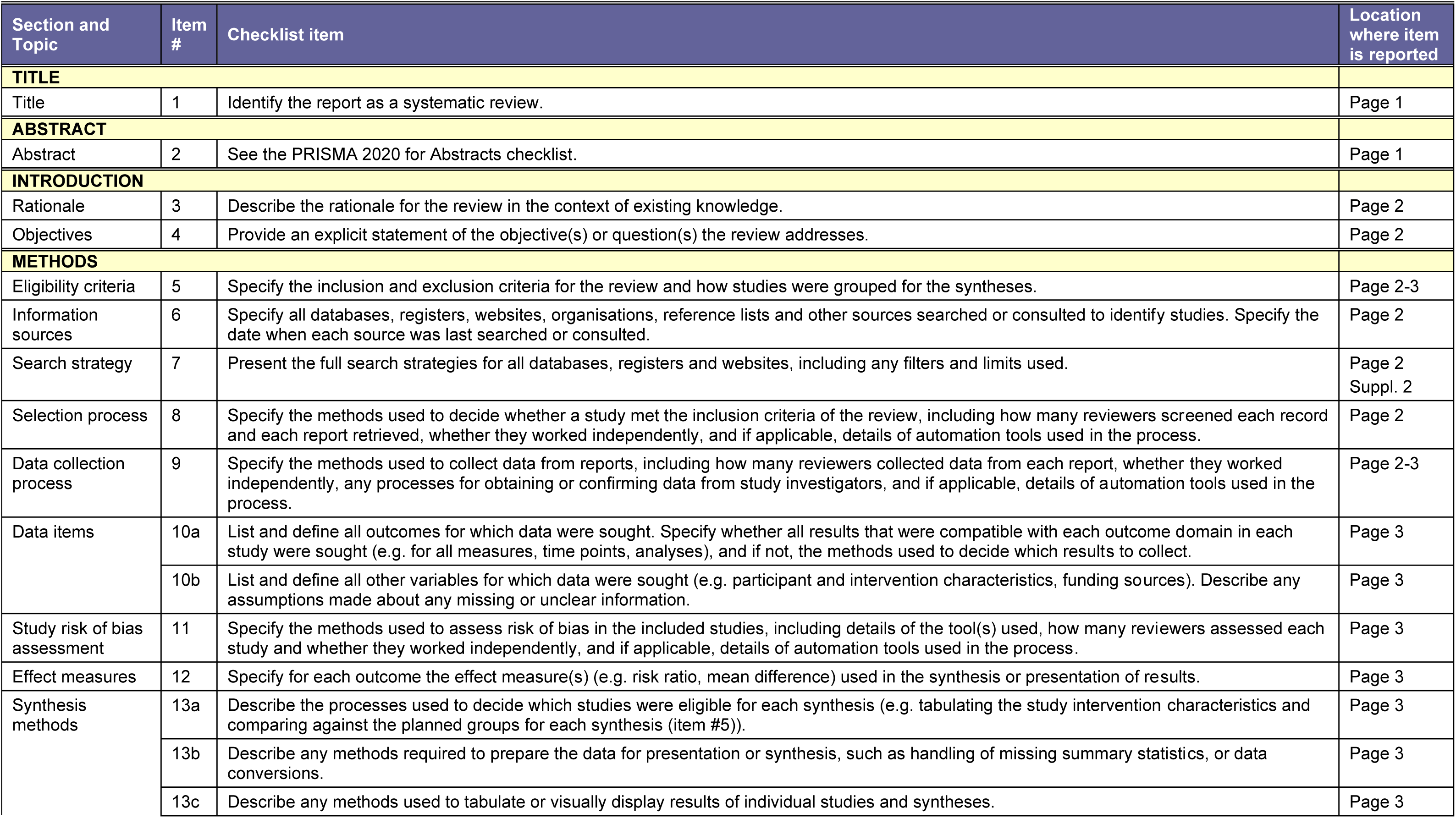
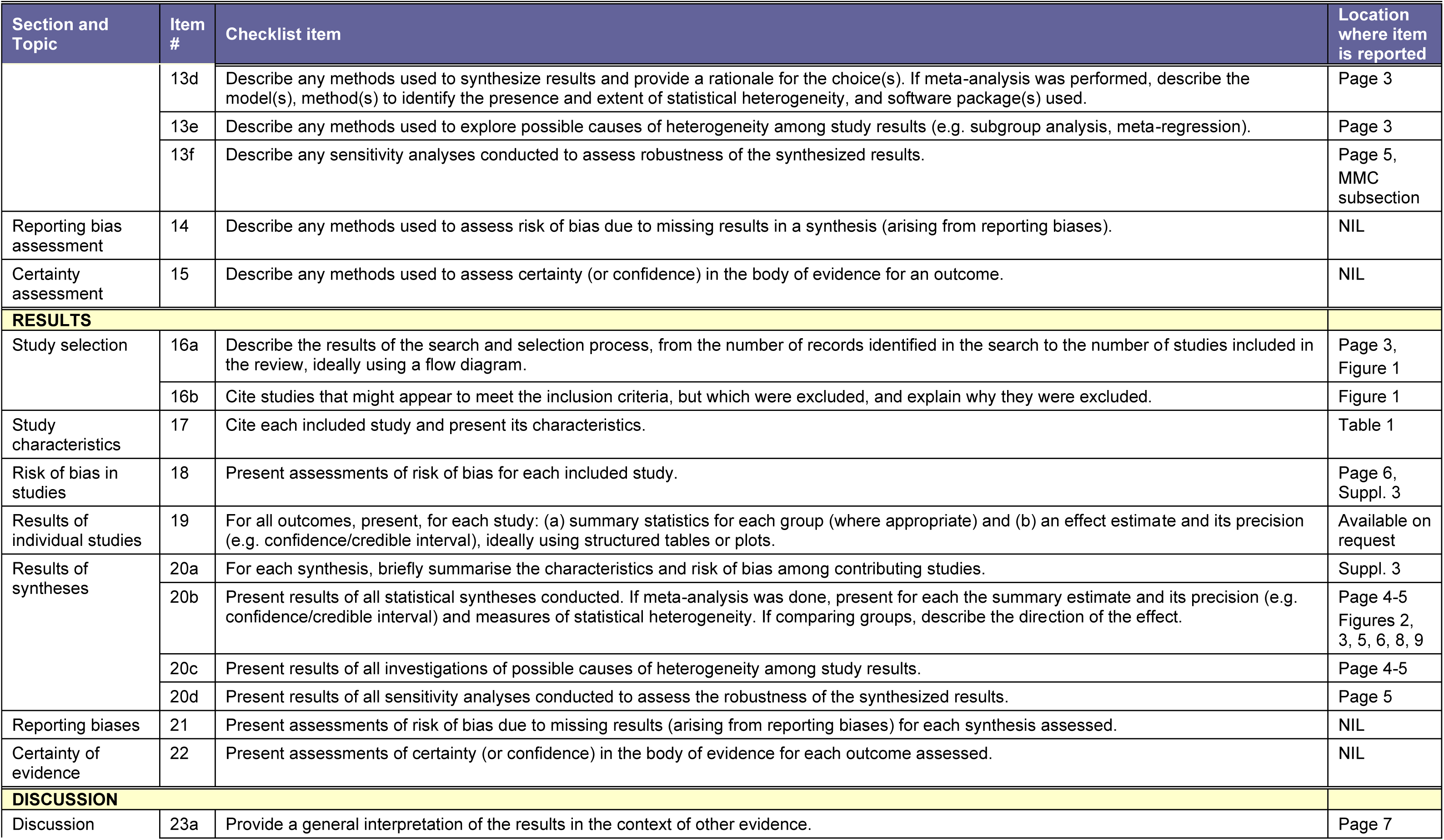
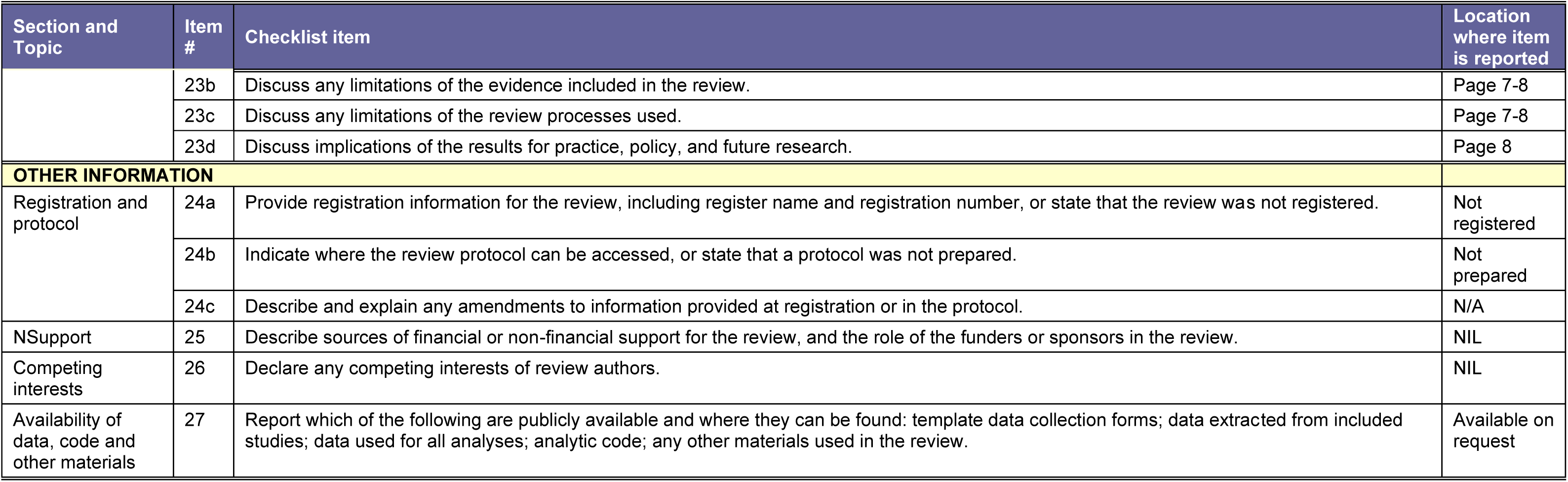
PRISMA checklist.

**Supplementary 2:**
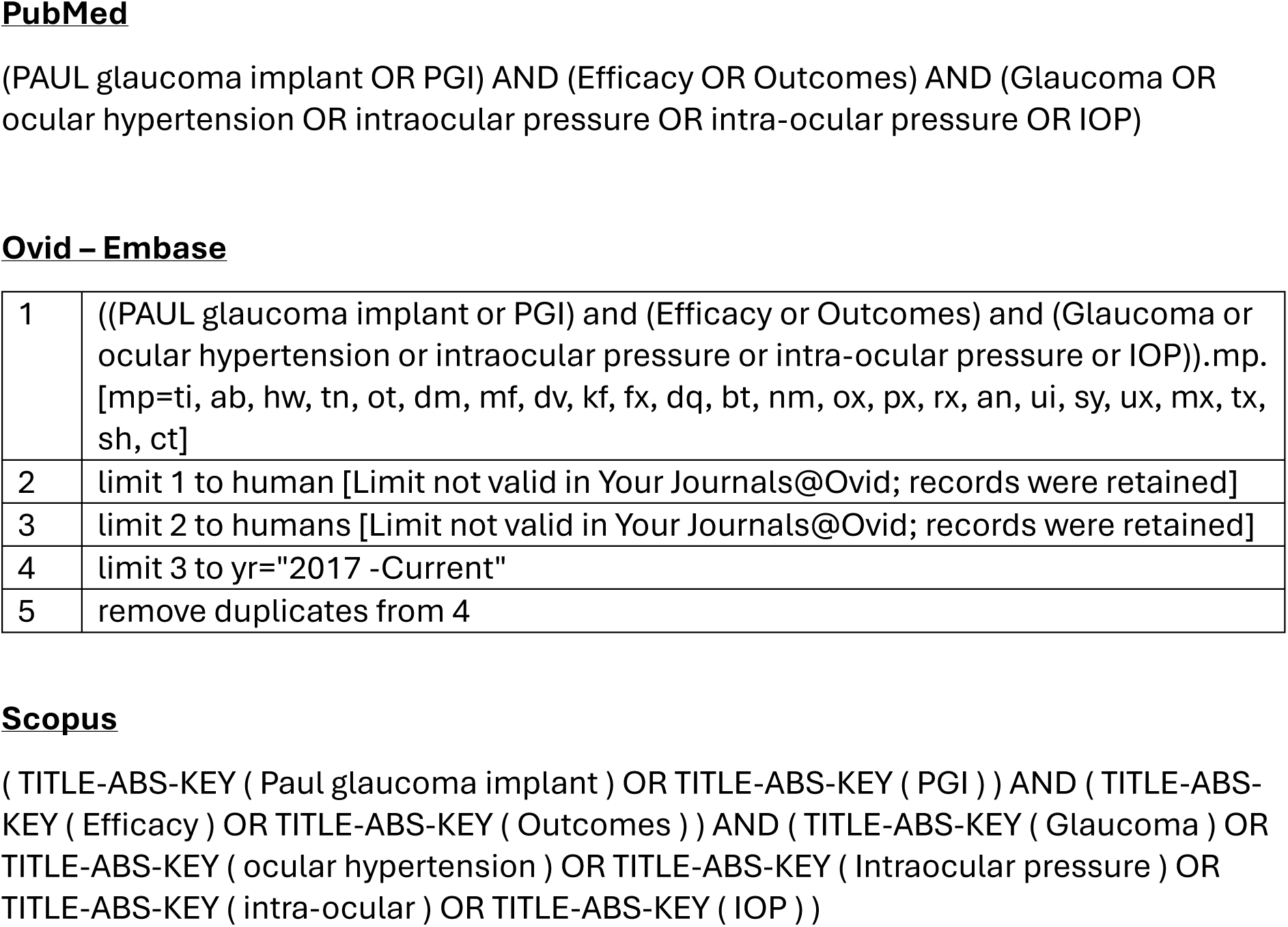
Search strategies.

**Supplementary 3:** Assessment of the quality of included evidence.

ROBUST-RCT for Randomised controlled trial (RCT)<sup>9</sup>
| Study, year | Item 1: Random sequence generation | Item 2: Allocation concealment | Item 3: Blinding of participants | Item 4: Blinding of healthcare providers | Item 5: Blinding of outcome assessors | Item 6: Outcome data not included in analysis |
| --- | --- | --- | --- | --- | --- | --- |
| Elhusseiny et al. 2025<sup>21</sup> | Definitely yes<br>Definitely low risk of bias | Probably no<br>Definitely low risk of bias | Definitely no<br>Definitely low risk of bias<br>(unblinding of participants very unlikely to have influenced outcome) | Definitely no<br>Probably low risk of bias<br>(unblinding of healthcare providers unlikely to have influenced outcome) | Definitely no<br>Probably low risk of bias<br>(outcome assessment involves objective outcomes) | 0% of data not analysed<br>Definitely low risk of bias |

Newcastle-Ottawa scale for non-RCTs<sup>8</sup>
| Study, year | Selection |  |  |  | Comparability | Outcome |  |  | Sum |
| --- | --- | --- | --- | --- | --- | --- | --- | --- | --- |
|  | Representativeness of exposed cohort | Selection of non-exposed cohort | Ascertainment of exposure | Demonstration that outcome of interest was not present at start of study | Comparability of cohorts on basis of design or analysis | Assessment of outcome | Was follow-up long enough for outcomes to occur | Adequacy of follow up of cohorts |  |
| **Adult** |  |  |  |  |  |  |  |  |  |
| Deubel et al. 2025<sup>29</sup> | * | N/A | * | * | N/A | * | * | * | 6 |
| Liegl et al. 2025<sup>30</sup> | * | N/A | * | * | N/A | * | * | * | 6 |
| Mendoza-Moreira et al. 2025<sup>20</sup> | * | * | * | * | * | * | * | - | 7 |
| Olgun and Karapapk et al. 2025<sup>31</sup> | * | * | * | * | * | * | * | - | 7 |
| Oliver-Gutierrez et al. 2025<sup>19</sup> | * | * | * | * | - | * | * | - | 6 |
| Weber et al. 2025<sup>32</sup> | * | N/A | * | * | N/A | * | * | - | 5 |
| Studsgaard et al. 2025<sup>33</sup> | * | N/A | * | * | N/A | * | * | * | 7 |
| Khodeiry et al. 2025<sup>34</sup> | * | N/A | * | * | N/A | * | * | * | 7 |
| Labay-Tejado et al. 2024<sup>35</sup> | * | N/A | * | * | N/A | * | * | * | 7 |
| Weber et al. 2024<sup>36</sup> | * | N/A | * | * | N/A | * | * | - | 6 |
| Berteloot et al. 2024<sup>18</sup> | * | * | * | * | * | * | * | - | 7 |
| Tan et al. 2024<sup>24</sup> | * | N/A | * | * | N/A | * | * | - | 5 |
| Karapapak and Olgun 2024<sup>17</sup> | * | * | * | * | * | * | * | - | 7 |
| Olgun and Karapapak 2024<sup>37</sup> | * | * | * | * | * | * | * | * | 8 |
| Richardson et al. 2024<sup>22</sup> | * | N/A | * | * | N/A | * | * | * | 6 |
| Hu et al. 2024<sup>14</sup> | * | N/A | * | * | N/A | * | * | - | 5 |
| Rusmayani et al. 2024<sup>38</sup> | * | N/A | * | * | N/A | * | * | - | 5 |
| Jose et al. 2022<sup>39</sup> | * | N/A | * | * | N/A | * | * | - | 5 |
| Vallabh 2022<sup>23</sup> | * | N/A | * | * | N/A | * | * | - | 5 |
| Koh et al. 2020<sup>40</sup> | * | N/A | * | * | N/A | * | * | * | 6 |
| **Paediatric** |  |  |  |  |  |  |  |  |  |
| Karapapak and Olgun 2024<sup>41</sup> | * | N/A | * | * | N/A | * | * | - | 5 |
| Mendoza-Moreira et al. 2024<sup>42</sup> | * | N/A | * | * | N/A | * | * | - | 5 |
| Vallabh et al. 2023<sup>43</sup> | * | N/A | * | * | N/A | * | * | - | 5 |

N/A if studies only have single cohort.

